# Evaluating Spatially Targeted HIV Interventions and Harm Reduction Services Among People Who Inject Drugs in a High-Burden Setting

**DOI:** 10.64898/2026.02.07.26345824

**Authors:** Jasmine Wang, Steven J. Clipman, Shruti H. Mehta, Aylur K. Srikrishnan, Shobha Mohapatra, Muniratnam S. Kumar, Gregory M. Lucas, Carl A. Latkin, Sunil S. Solomon, Amy Wesolowski

**Affiliations:** Department of Epidemiology, Johns Hopkins Bloomberg School of Public Health, Baltimore, MD, USA; Division of Infectious Diseases, Department of Medicine, Johns Hopkins University School of Medicine, Baltimore, MD, USA; YR Gaitonde Centre for AIDS Research and Education, Chennai, India

## Abstract

People who inject drugs (PWID) in India continue to experience high HIV incidence while coverage of HIV and harm reduction services within this population remains suboptimal in many settings, highlighting the need to identify novel service delivery points. To evaluate the effectiveness of spatially focused upscaling of interventions at observed venues where PWID injected drugs together, we developed an individual-based dynamic transmission model of HIV informed by detailed injection network, service engagement, and injection venue attendance data collected in a sociometric study of PWID (n = 2512) in New Delhi, India. HIV incidence was simulated for different spatial targeting strategies and with increasing service coverage at injection venues according to UNAIDS/UNODC goals. We identified significant decreases in predicted HIV incidence when deploying interventions at frequently visited injection venues (from 6.8 cases/100 person-years to 2.7/100PY for full service coverage at the most-visited venue, and further down to 1.3/100PY for 12 most-visited venues). Prioritizing the most visited venues stratified by spatial clusters provided services to a larger number of individuals versus prioritizing the overall most visited venues, suggesting that service expansion at venues that are spatially distinct with minimal population overlap has a slightly larger impact on reducing HIV incidence.

## Introduction

People who inject drugs (PWID) account for a disproportionate burden of human immunodeficiency virus (HIV) globally and are contributing to some of the fastest-growing epidemics in an era of continued progress toward HIV control[1–3]. A high HIV burden, between 6% to 10% prevalence, has been reported among PWID in India[4] with significant regional variance[5] and incidence rates higher by an order of 10 folds than other vulnerable populations such as female sex workers and men who have sex with men[6]. Partially contributing to this high prevalence is low engagement in harm reduction services among PWID in India[7]. For example, even after an expansion in 2020-2021, medication for opioid use (MOUD) centers were estimated to cover less than 10% of local PWID in most states of India[8], far below the goal of 50% coverage for people with opioid dependence by 2025 from the Global AIDS Strategy[9, 10].

Factors that drove suboptimal coverage include criminalization of drug use, limited access to health care services, and economic instabilities resulting in many barriers to accessing care[11, 12]. Although efforts continue to expand MOUD services alongside syringe service programs (SSP) have occurred in recent years, to date, there has been little impact on reducing HIV burden in this group[4].

One of the challenges in the delivery of harm reduction services to PWID is how to optimize access and engagement when limited resources/programs are available.

Commonly used strategies, such as decentralized service provision and peer-delivered harm reduction, can be effective but are often limited by their ability to reach either enough individuals or those at the highest risk[13]. The Joint United Nations Programme on HIV/AIDS (UNAIDS) 2025 targets for service coverage specify that 95% of individuals living with HIV should be diagnosed and that 95% of those diagnosed should be on sustained antiretroviral treatment (ART)[10]; United Nations Office on Drugs and Crime (UNODC) goal for SSP coverage is 200 sterile needles and syringes per PWID per year[10], and 50% of individuals with opioid dependence should get access to MOUD according to the Global AIDS Strategy[10]. These goals require service delivery strategies that consistently cover a high percentage of the PWID population. Spatially targeting prevention programs have been proposed as a means to maximize efficiency since clusters of HIV cases tend to be heterogeneously distributed geographically[14].

Previously, we explored the spatial and network factors associated with high-risk injection practices and HIV transmission among a cohort of PWID in New Delhi, India[15, 16]. We found that while needle-sharing partnerships were dynamic, there was strong geographic clustering of risk behaviors and HIV transmission by injection venues. Risk spaces other than PWID residences have been previously identified as loci of concentrated high-risk activities[17], emphasizing the necessity to incorporate spatial factors into PWID network analyses. Understanding the heterogeneity and spatial distribution of PWID risk behaviors and linkage-to-care is indispensable to more efficiently deploy interventions to population clusters that are at high risk or disproportionately contributing to onward transmission. At the same time, leveraging the number of unique individuals at each injection venue and the geographical proximity of these venues could help optimize the deployment of any injection venue-focused strategies. Thus, we aim to explicitly incorporate injection venues as the primary sites for PWID harm reduction and HIV clinical service delivery in an HIV transmission model to a) project changes in HIV incidence by this spatially focused strategy and b) explore how different orders of prioritizing venues for service delivery translate to intervention effectiveness.

Here, we leverage a highly granular and comprehensive cross-sectional dataset of PWID with both their social (injection partnership) and spatial (injection venue attendance) networks explicitly mapped. We analyzed the injection venue attendance of a network-based cohort of PWID in New Delhi, India, and estimated their spatial patterns of injection-related risk behaviors and service access by injection venue attendance. We developed an individual-based network model of HIV transmission among a population of PWID with similar characteristics to the New Delhi cohort to assess spatial targeting strategies for service delivery. Injection partnership, venue attendance, demographics, risk behaviors, and baseline service engagements were informed by analyses of the empirical cohort data and dynamically modeled. We then evaluated the impact of scaling up interventions at injection venues, which increased the probability of service engagement for individuals who frequent these venues. We compared two different deployment strategies of either prioritizing venues for service expansion based on a) the number of PWID who frequent each venue (population-weighted) or b) a population-weighted strategy stratified by geographical locations. In all simulations, we evaluated the impacts of a range of service coverage levels and the number of venues with enhanced services on changes in HIV incidence in the simulated population compared to baseline, with service coverage as the status quo.

## Results

### Spatial patterns of PWID risky behaviors were heterogeneous and concentrated at a small number of injection venues

Based on responses from 2512 participants, a total of 110 injection venues with >10 unique visitors were identified at baseline (Figure 1). Participants reported frequenting a mean of 4.4 venues (IQR = [2,6]). To explore possible differential injection venue attendance by demographic or behavioral characteristics, we analyzed several variables reflecting injection-related risk behaviors and the uptake of harm reduction or clinical services stratified by whether the participant visited the top 5 populated venues, which we considered as popular venues amongst our cohort. These top 5 venues were cumulatively visited by ∼66% of our baseline PWID cohort. Participants who reported attendance at popular venues tended to be younger and initiated injection drug use at a younger age; these individuals also reported having a larger number of injection partners and visiting more injection venues (Table 1). In terms of risk behaviors, individuals visiting these popular venues had higher injection frequencies and reported that a higher proportion of their injections were with others. They were also less likely to have ever been tested for HIV while also more likely to be living with HIV. Harm reduction service engagement was less clear by injection venue as participants frequenting popular venues were less likely to access MOUD but more likely to access SSP (all p < 0.05). When considering individual injection venue populations and behavioral characteristics of their visitors, however, there was no significant linear relationship between venue population and variables commonly related to HIV transmission (Figure S1).

**Figure 1.**
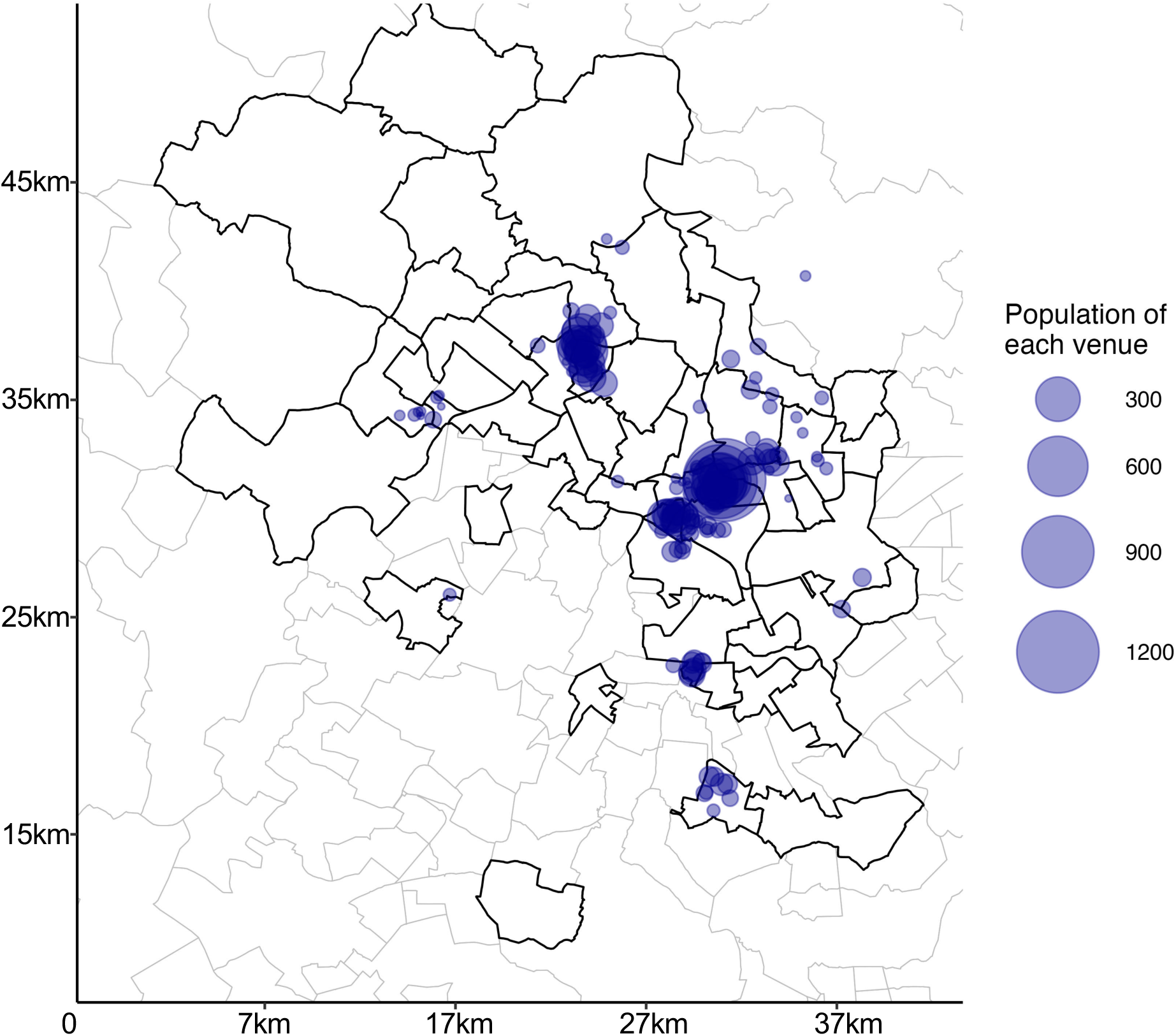
Postal code map of residence and injection venues of PWID surveyed within the New Delhi cohort. Postal code regions with solid boundaries are residences of participants, and blue dots are locations where at least 10 participants reported injecting drugs (injection venues) with sizes scaled to their visitor populations.

**Table 1.** PWID behaviors and service access stratified by popular venue attendance. Characteristics of a total of 2512 participants were compared by whether they visited the top 5 most frequently visited injection venues. Two-sided Wilcoxon rank sum tests for continuous variables and two-sided Pearson’s Chi-squared tests for categorical variables were conducted. MOUD = medication of opioid use disorder; SSP = syringe service program.

|  | Participants who visited the top 5 populated venues, N = 1,662<sup>1</sup> | Participants who did not visit the top 5 populated venues, N = 850<sup>1</sup> | p-value<sup>2</sup> |
| --- | --- | --- | --- |
| Age | 29 (22, 32) | 31 (22, 36) | 1.3e-5 |
| Age of first drug injection | 22 (17, 25) | 23 (18, 27) | <1e-5 |
| Monthly injection frequency | 68 (30, 90) | 56 (24, 90) | <1e-5 |
| Proportion of injections with other individuals | 0.56 (0.30, 0.84) | 0.53 (0.27, 0.75) | 0.023 |
| Proportion of injections where syringes were shared | 0.30 (0.00, 0.67) | 0.31 (0.00, 0.75) | 0.59 |
| Ever tested for HIV | 712 (43%) | 492 (58%) | <1e-5 |
| Living with HIV | 690 (41%) | 239 (28%) | <1e-5 |
| Injection partners in the past month | 3 (2, 4) | 3 (2, 3) | 1.8e-4 |
| Number of injection venues visited | 5.0 (2.0, 7.0) | 3.2 (1.0, 4.0) | <1e-5 |
| Days accessed MOUD past 6 months | 38 (0, 180) | 69 (0, 180) | <1e-5 |
| Days accessed SSP past 6 months | 19 (0, 180) | 9 (0, 180) | 4.8e-5 |

<sup>1</sup> Mean (IQR); n (%)
<sup>2</sup> Wilcoxon rank sum test; Pearson's Chi-squared test

### Injection venues were spatially clustered, allowing for spatial targeting of interventions

Three spatial clusters of injection venues were identified by matching venues to postal code areas of New Delhi and calculating the total number of injection venues within each postal code area (Figure 2a). We compared the number of injection venues in each postal code area versus their neighboring postal codes and identified three geographically disparate areas (with postal codes as units) that contained 35, 89, and 8 unique injection venues and spanned an area of 72, 148, and 11 square kilometers, respectively (see Figure 2a).

**Figure 2.**
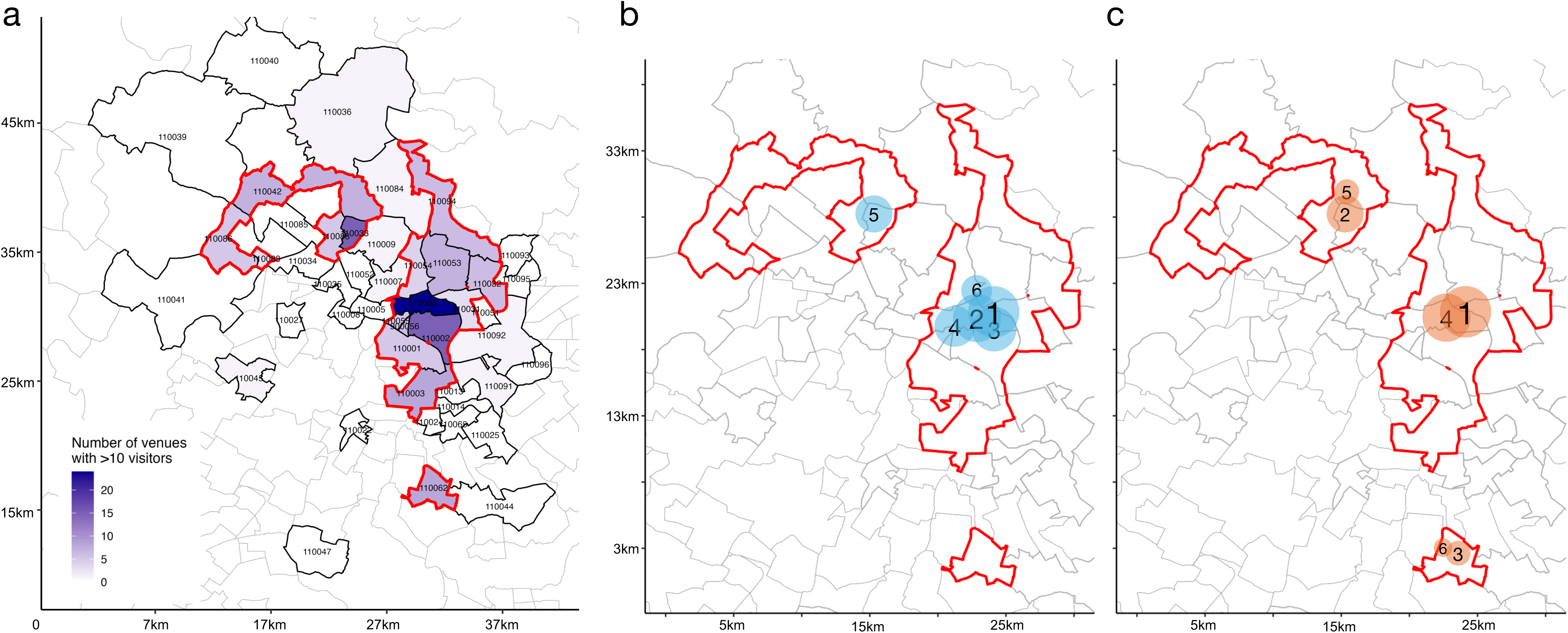
Postal code location by number of injection venues and simulated spatial expansion strategy. a) Postal code regions are shaded based on the number of injection venues. When simulating spatial intervention expansions, we considered three clusters of injection venues (outlined in red) and the corresponding postal codes. Zoomed-in maps of these venue clusters were shown with example orders of service delivery based on the two strategies, including the b) population-weighted approach and c) spatially-stratified approach. In both instances, the size of the circles represents the relative population sizes of venues, and the number corresponds to the rank order. Locations of venues on maps were approximate. For the population-weighted approach, the order of service delivery was determined by the venue population. In the spatially-stratified approach, the order of service delivery was first ranked by spatial clusters of the venues to provide services in geographically disparate locations and then by venue population.

PWID residences were often near their reported injection venues, although some of the most popular venues were also frequented by PWID who lived further away (Figure S2-3). Despite the overlapping residence locations of PWID from all 3 clusters, each cluster included at least one unique postal code area of residence, and over 75% of resident PWID frequented that cluster. Higher percentages of PWID living in the northwest postal code areas of New Delhi visited injection venues in Spatial Cluster #1. Most PWID living in the central postal code areas visited venues in Cluster #2, as did most PWID from the peripheral postal code areas. Cluster #3 was one single postal code area, but its venues were almost exclusively frequented by PWID living in that postal code area, who did not visit any other venues in Clusters #1 or #2.

By adding up unique participants who reported visiting each venue in their survey responses, we compared the numbers of unique PWID covered by scaling interventions spatially: we considered two different approaches to expand services to injection venues (Figure 2b, c). The population-weighted approach sequentially increased service coverage at overall popular venues frequented by the most individuals, while the spatially-stratified approach sequentially covered the most popular venues by each geographical cluster (see Methods). As shown in Figure 3, deploying services in the order of the spatially-stratified approach consistently reached a larger population than the population-weighted approach when covering no more than 9 injection venues. The relationship was flipped when more than 9 venues were covered since the spatially-stratified approach included more venues that were in remote locations. Even when compared to a Greedy Algorithm selection process, where services are expanded to maximize the number of unique number of PWID with improved care, the spatial-stratified approach yielded smaller gaps in PWID coverage from the Greedy Algorithm for the first several venues covered (Figure S4).

**Figure 3.**
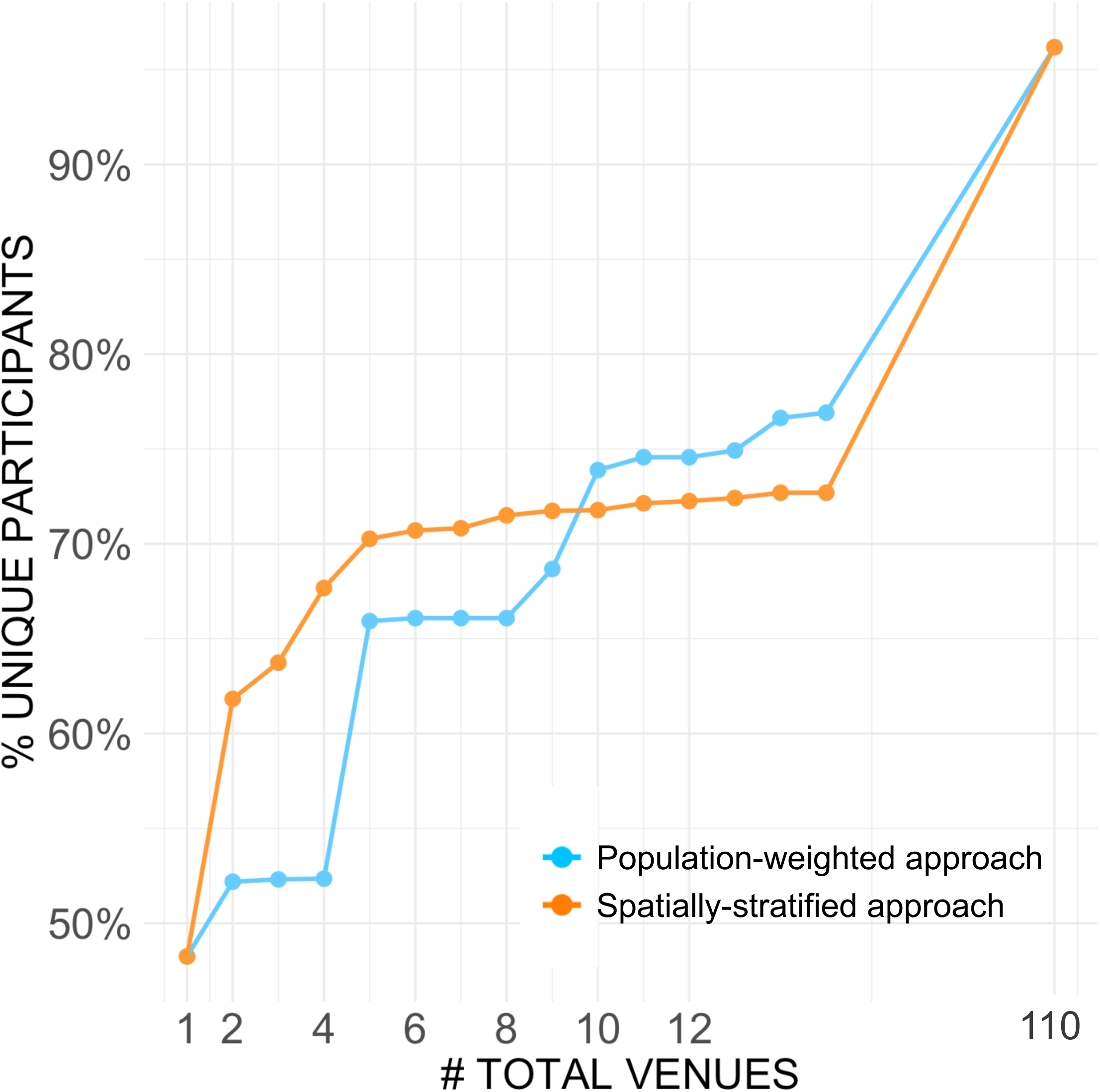
Cumulative unique PWID covered by incrementally adding venues to target for service delivery. Percentage of total unique individuals covered by interventions at injection venues based on two service expansion strategies. The x-axis showed the total number of venues covered based on either strategy, from 1 to all 110 venues with >10 individuals.

### Model simulations with service coverage ladder and two expansion approaches

We leveraged survey data from the cohort in India to calculate estimates of injection partnerships, injection venue attendance, risk behaviors, and service access. We calculated venue-specific risk behaviors and service access parameters for the model that would reflect the characteristics of the sub-population who visited a particular injection venue (Table S1). A model schematic (Figure S5), table of parameters (Table S2), and ERGM diagnostics and goodness-of-fit (Figure S6) are provided in Supplementary Materials. All simulations started with the same prevalence of people living with HIV observed in the cohort (37% prevalence) for all locations. We then explored scenarios with expansions in both 1) the number of injection venues to scale up service coverage for PWID who frequented the venue and 2) the coverage levels (50%/75%/100% to UNAID/UNODC goals for each service). For each scenario, we ran 50 simulations to account for stochasticity. We present the mean HIV incidence 2 years after service deployment to quantify the effectiveness of combinations of service coverage and different expansion strategies.

Without scaling up any interventions, the HIV incidence of the simulated population stabilized at 6.8 cases per 100 person-years, with a 95% confidence interval of [6.5, 7.2]. When increased coverage of a package of services (SSP, MOUD, HIV testing, and ART prescription) was simulated, unsurprisingly, HIV incidence decreased as more injection venues were covered (Figure 4). Taking the spatially-stratified approach as an example (Figure 4a), when 50% of all UNAID/UNODC goals were reached, the resulting HIV incidence decreased to 3.8 per 100PY [3.4, 4.2] when interventions were expanded to only the most populated venue; HIV incidence further dropped to 2.9 per 100PY [2.6, 3.3] when 12 out of 110 injection venues were covered. When 100% of all UNAID/UNODC goals were reached, HIV incidence was 2.7 per PY [2.5, 3.0] when services were scaled up at the most populated venue and could go down to 1.3 per 100PY [1.1, 1.5] when 12 venues were covered. The population-weighted venue expansion approach led to similar results (Figure 4b). Across the range of service coverage combinations, scaling SSP coverage resulted in the largest marginal decrease in HIV incidence. Improving SSP coverage closer to the UNODC goals (50% to 75%, or 75% to 100%) decreased HIV incidence by ∼10%.

**Figure 4.**
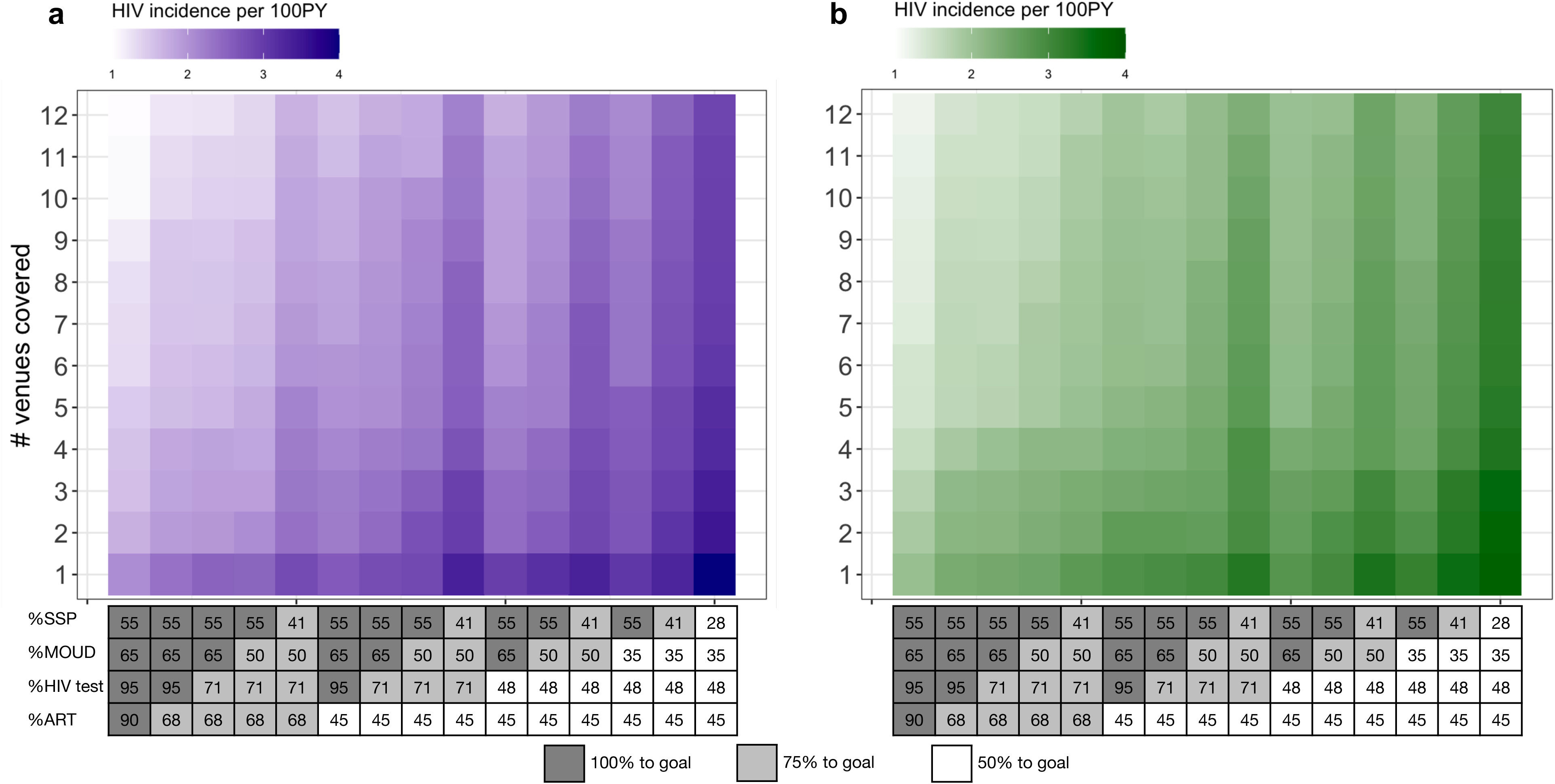
HIV incidence per 100 person-years two years after intervention expansion, with an increase in service coverage per injection venue and number of venues covered using two targeting approaches. We considered both the a) spatially-stratified b) population-weighted approaches when ranking injection venues to increase coverage. The ladder of increased coverage for each service is shown in the bottom table, with the value in each cell representing the percentage of service coverage at a venue. ART = antiretroviral therapy; MOUD = medication for opioid use disorder; SSP = syringe service program.

To further explore the effectiveness of the two expansion approaches, we first analyzed the number of unique individuals covered when the number of covered venues increased, using SSP as an example (Figure S7). The overall marginal advantage of the two strategies closely reflects the number of unique PWID covered in Figure 3. When SSP is scaled up, regardless of the target coverage (% to goal), the spatially-stratified approach yielded significantly more unique individuals covered by SSP compared to the population-weighted approach. When services were expanded to more than 4 venues, the differences between the two strategies were negligible (Figure S7).

Finally, we evaluated how the two expansion strategies impacted the decrease in HIV incidence within the simulated population (Figure 5). For simplicity, here we only compared three levels of overall interventions: low/medium/high levels, as in all services (SSP, MOUD, HIV testing, and ART prescription) being all scaled up to 50%/75%/100% of their respective goals. As expected, HIV incidence decreased as service coverage improved at more injection venues. Across all three coverage levels, service delivery to the most populated injection venues led to the largest marginal decrease in HIV incidence, followed by decreases at slower rates when interventions were expanded to less populated venues. Scaling up all interventions in all 110 venues with >10 visitors decreased HIV incidence to 2.6 per 100PY [2.4, 2.9] for the low service coverage level and 0.5 per 100PY [0.4, 0.6] for the high coverage level. In terms of changes in HIV incidence per venue covered, we estimated a decrease of 4.1 per 100PY per venue in HIV incidence when covering the first venue, 0.82/100PY/venue when covering 6 venues, and 0.04/100PY/venue when covering all 110 venues. Unlike the unique numbers of individuals who benefited from either expansion strategy (Figure 3), the relative difference in HIV incidence between the two expansion strategies was smaller and less statistically significant (Figure 5). Expanding services from 2 to 4 total venues yielded a slight marginal advantage for the spatially-stratified approach, but the difference was not statistically significant. The largest marginal decrease in HIV incidence when adopting the spatially-stratified approach was when the 2 top venues were targeted, with an additional 0.27 per 100PY decrease in HIV incidence. When services were expanded to more than 4 venues, there was no substantial advantage by either expansion strategy in this specific PWID population. While neither service strategy was able to reach the entire PWID cohort even with 12 venues being deployed interventions, the individuals who were not reached in our simulation showed lower risks of HIV transmission, compared to those who were reached (Table S3).

**Figure 5.**
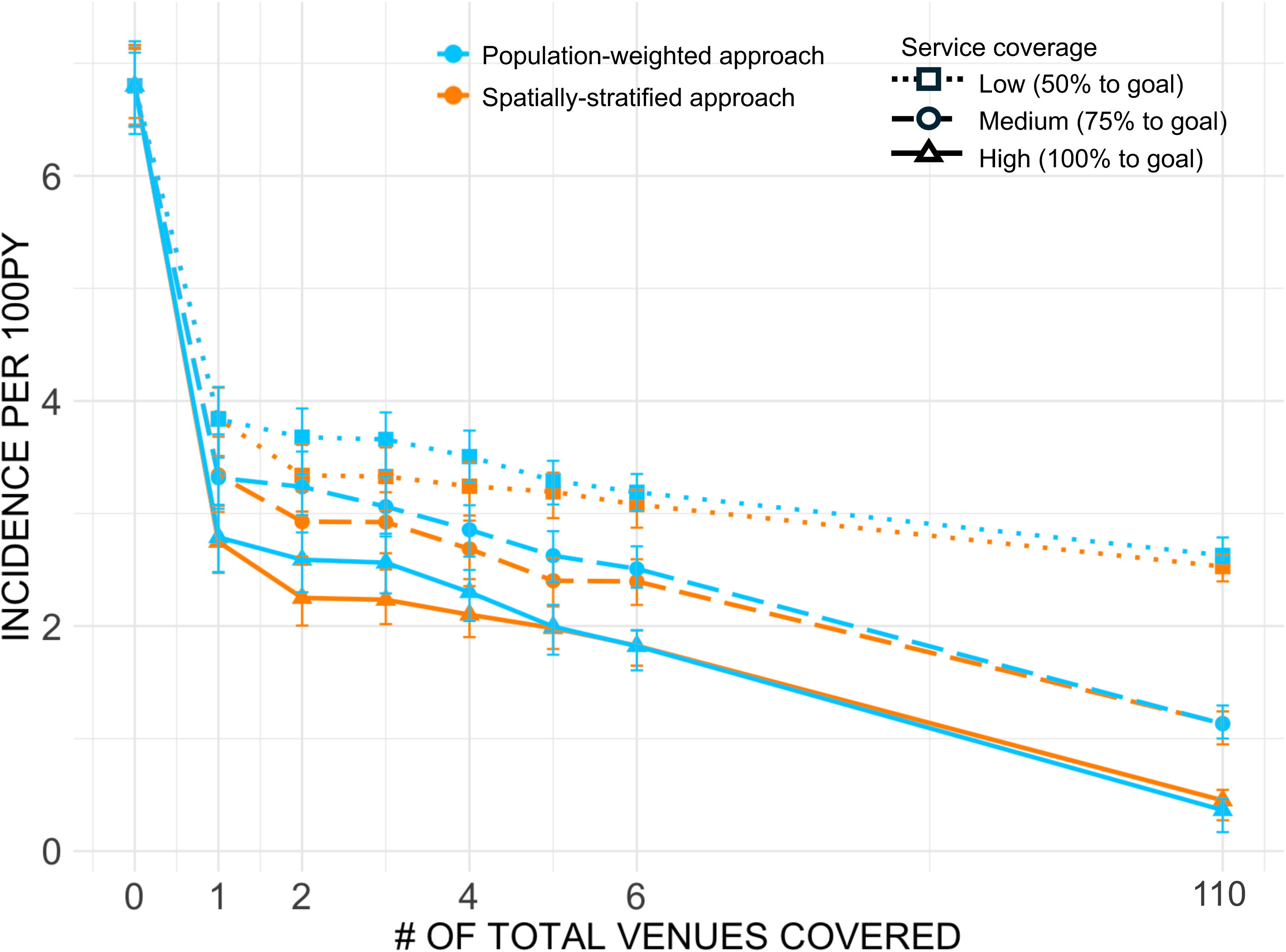
HIV incidence per 100 person-years two years after intervention. Three levels of upscaled SSP, MOUD, HIV testing, and treatment altogether were simulated at 0 to 110 total injection venues by different venue priority ranking approaches for service deployment. Points represent mean values of 50 stochastic simulations, and error bars were upper and lower bounds of 95% confidence interval of simulated incidence values.

Given the highly skewed visitor counts among injection venues with the most popular venue being frequented by 48.5% of our PWID cohort, we considered a counter-factual scenario where the most popular venue did not exist and thus interventions could not be scaled up. This scenario led to fewer individuals being reached by targeting a few injection venues, as well as a smaller difference in the unique numbers of PWID covered by the two service expansion strategies, with the spatially-stratified strategy reaching more individuals until 8 injection venues were covered (Figure S8). Like Figure 5, Figure S9 demonstrates the decrease in HIV incidence if various levels of upscaled services were deployed to an increasing number of injection venues, assuming the most popular venue identified from our data did not exist. Decreases in HIV incidence by upscaled interventions were smaller when injection venue attendance was more scattered; for example, HIV incidence ranged from 2.3 to 3.5 cases per 100PY when 6 venues were covered by either service deployment approach (Figure S9) compared to 1.9-3.2 cases per 100PY with the most popular venue being covered first (Figure 5). When only few venues (2-3) were covered, the spatially-stratified approach resulted in significantly lower HIV incidence (p < 0.05) compared to the population-weighted approach, but the difference became negligible when more than 3 venues were covered.

## Discussion

Expanding services in a spatially focused manner can improve the effectiveness of harm reduction and HIV-care interventions among PWID, given the spatial heterogeneity in risk behaviors and HIV infections[14, 18]. However, few studies can quantify the possible magnitude of this effect since it requires empirical information on socio-spatial PWID networks coupled with risk behaviors and service uptake. Using rich behavioral data and an individual-based model parameterized to these data, we investigated the potential impact of service expansion to injection venues. Our results showed that higher HIV prevalence and risky drug-injection correlates, including younger age, low service engagement, higher injection frequency, and a larger number of injection partnerships, were strongly associated with popular injection venue attendance. This finding indicates significant spatial concentrations of vulnerabilities, risk behaviors, and infections[14]; these hotspots can thus become origins of potential HIV and other infectious disease (e.g., Hepatitis C) outbreaks or the most effective locations to deploy scaled-up services, emphasizing the realistic benefit of a spatial model of intervention deployment. Three disparate clusters of highly populated injection venues were identified, where individuals often frequented a venue near their reported residence, suggesting that these clusters may be stable over time. Two different spatial strategies of venue priorities were investigated. Our simulation results showed that HIV incidence had the largest decrease when services were upscaled in the most populated venue, achieving a decrease from a baseline of 6.8 per 100PY to 2.7 per 100PY if 100% of UNAID/UNODC goals were reached. The marginal reduction in HIV incidence decreased as coverage increased across more venues (down to 1.3 per 100PY when 12 venues were covered and 0.5 per 100PY when all 110 venues were covered). This was related to the high number of individuals frequenting the most popular injection venue and the decreasing number of unique PWID who exclusively frequented less popular venues. We also simulated a hypothetical ladder of services at each expanded site, with the idea of an integrated service delivery model (MOUD, SSP, HIV testing, and ART prescription) similar to what has been recommended by UNAIDS and is currently in use across multiple cities in India[19]. Considering the effectiveness of each individual service, SSP yielded the largest decrease in HIV incidence, given the equivalent magnitude of service expansion. Finally, HIV incidence from our simulations 2 years after scaled-up interventions did not significantly differentiate the effectiveness of the two spatial targeting strategies. This was likely since the spatial network reported by our cohort was highly skewed towards the most popular injection venue, leaving less space for significant gaps in HIV incidence drop between the two ranking approaches when more venues are covered. Individuals who visited the more populated venues also had more injection partners and engaged in risky injection behaviors more frequently, resulting in another layer of indirect effect on HIV transmission with the population-weighted approach. As a result, although the spatially-stratified strategy covered more unique PWID faster than the population-weighted strategy, the latter could reach more high-risk individuals faster, given the association between popular venue attendance and HIV transmission risk. Our conclusion emphasized the nuanced and correlated population-and individual-level contributors to be considered when designing service expansion interventions specific to the socio-spatial context of the PWID population of interest.

Our results were based on several key assumptions. First, we based our conclusions on cross-sectional data of injection venue attendance of our PWID cohort, thus ignoring the evolution of venues, including the potential formation of new locations and dissolution of existing ones. The difference in service delivery efficiency between the two strategies we tested can change in either direction, for example, if the area hosting the largest venues was evacuated or if smaller venues were more likely to dissolve in time. We also assumed that providing services at a venue would directly lead to higher service engagement rates. This tends to be true in settings with low service coverage, such as India, where service availability, instead of service uptake or awareness, is likely the bottleneck for increasing overall service engagement[8].

However, in other settings where service uptake is less related to access, spatial expansion of services may not be as effective. Further, we used sociometric data from a single cohort, which has a highly skewed venue attendance distribution. If venue attendance were more evenly dispersed in other networks of PWID, then the two spatial expansion strategies would likely show a larger difference in HIV incidence reduction.

Similarly, while our data did not show a significant pattern in the heterogeneity of individual service engagement across injection venues (e.g., smaller venues having significantly lower SSP access rates), PWID in other geographical contexts may form clusters that have specific needs for one or more harm reduction or clinical services. Another aspect that limits the generalizability of our study was the strongly coupled spatial and social networks in our PWID cohort. Here, we prioritized the spatial component to inform probabilistic formation of social/injection partner ties in our PWID network. This way of simulating the dynamics of a PWID network may be less applicable in other PWID populations if injection ties are driven by factors other than injection venue attendance. Further, we did not include concurrent sexual transmission amongst PWID, which has been reported in other PWID studies[20]. Our cohort reported very few IDU sexual partnerships, which could indicate separate drug-use and sexual networks for PWID. Finally, our model was highly dependent upon detailed sociometric data, including uniquely identified visitors at injection venues, to inform the two strategies. These data are likely unavailable in other settings; however, general estimates of injection venue population sizes could be informed by active and systematic mappings of suspected injection locations followed by interview verifications, or could be captured as part of routine surveillance efforts [21]. These methods are easier to replicate and could also potentially capture the spatial heterogeneity of PWID populations of interest, especially highly clustered spatial networks; our model could then be readily applied to quantify venue-specific risks and intervention effectiveness.

Our study leveraged empirical socio-spatial network data of a PWID cohort from an LMIC setting to quantify the effect of spatially improving the delivery of HIV care and harm reduction services with a transmission model. Existing service delivery points at facilities such as government hospitals can be viewed as being ‘overly medicalized’ by the community[22], while the decentralized service delivery to injection venues can not only improve spatial proximity between service provision and where PWID naturally gather but can also reach high-risk PWID who are less likely to engage in facility-based services. This strategy is operationally feasible as trained outreach workers and mobile vans are able to access these identified venues and deliver services at the field-level.

Injection venues, especially those frequented by PWID living nearby, as in our cohort, are likely more stable than the drug-use dyadic relationships from PWID social networks that can dissolve and reform more randomly. Furthermore, these naturally formed injection venues can initiate chain effects; for example, individuals with access to SSP could act as secondary exchangers of clean syringes at these injection venues to further and indirectly increase the efficacy of upscaled interventions at a limited number of locations. Analysis of injection venues was able to identify high-risk HIV clusters within our cohort[15] while previous work analyzing urban PWID residential areas could not[23]. We evaluated the direct reduction in HIV incidence upon reaching the UNAIDS/UNODC coverage goals to different extents at these injection venues.

Although the strategy of prioritizing venues by a specific order would depend on the network composition and characteristics of the PWID population of interest, spatial approaches are likely to reach maximal effectiveness and consistent service engagement amongst PWID.

## Methods

### Study overview

Our study used empirical data from the baseline cohort of The Spatial Network Study, an ongoing longitudinal cohort of PWID in New Delhi, India, since 2017. All participants provided written informed consent. Details about the study population and recruitment have been previously published[15, 16]. The model was informed by survey data from 2512 participants who had their baseline visits between November 2017 and July 2019. Variables used included basic demographic characteristics (age, gender, etc.), injection venue(s) visited, drug-use behaviors including frequency of injection, injection with others, syringe sharing, service access rates including MOUD and SSP, HIV testing, and ART adherence. We also used test results from biospecimens, including HIV antibody titer and, if positive, HIV RNA level. As part of the study, each participant was asked to provide information about partners they had injected drugs with more than once in the past month. Unique referral cards were given to the participant for each injection partner nominated; if the named injection partner came back with the referral card, a network tie would be recorded. As a result, a sociometric network was built from 10 initial index participants and 2502 subsequent recruits. An additional list of confirmed deaths, migrations, and suspected deaths with corresponding participant IDs was used to estimate the mortality rate of the cohort. The study protocol was approved by institutional review boards at Johns Hopkins Medicine (IRB00110421) and the YR Gaitonde Centre for AIDS Research and Education in India (YRG292). All participants provided written informed consent.

### Network model structure

We adapted and extended the R package EpiModel modeling framework (Version 2.4.0)[24] to build a dynamic network-based model of HIV transmission. We simulated a population like the cohort, with 2512 individuals with the same demographic characteristics and similar sociospatial networks as in their baseline surveys. Model parameters were informed by cross-sectional analyses of the survey data (Table S2), when possible, as well as estimates from existing literature (details about the data source, uncertainty, and calibration of parameter values can be found in the Supplementary Material). A temporal exponential random graph model (ERGM) was used to simulate the dynamic and heterogeneous nature of injection partnerships, where nodes were individuals and edges/ties represented injection partnerships. We estimated an ERGM with a) network degrees associated with each node’s HIV infection status, number of injection venues they visited, and whether they were an index node, and b) assortative mixing associated with nodal homophily of popular venues both nodes visited and their HIV infection status. At each time point, individuals could enter the population and join the injection network or leave the population permanently based on an all-cause mortality rate specific to this cohort. Each individual newly entering the simulated population was assigned several injection venues to frequent based on a distribution of the number of venues visited by the surveyed cohort; the exact venues visited were randomly sampled with the probability ratio proportional to the population of each venue. Stochasticity was incorporated into the model using distributions of demographic and drug-use characteristics (see Table S2) for new PWID entering the population, venue-specific service engagement where the probability of engagement was informed by the data (Table S1) or simulated increases in service attendance, and the probabilistic formation and dissolution of social and spatial ties at each time point, with the dissolution rate from survey data. Simulations were run for 2 years after additional service expansion at injection venues (at year 1).

### Simulating disease transmission and service uptake

Given that our data mainly focused on details of the drug injection network compared to the IDU sexual network, we focused on parenteral transmission through sharing injection paraphernalia among PWID, as equipment sharing was identified as a substantial contributor to PWID morbidity and mortality globally[5]. The baseline HIV infection prevalence (defined by detectable HIV antibody titer) within the initial population was 37%[15]. Transmission can occur when an individual living with HIV (the ego) injects drugs with their network member(s) or vice versa. The number of injection events done with others was determined by the ego’s monthly injection frequency and whether the ego had access to MOUD that could decrease their frequency of drug use, multiplied by a proportion of injections shared with others by the ego (informed from cohort data). The injection dyad was more likely to inject together if they went to at least one common injection venue. A base probability of syringe sharing was applied to each injection partnership and proportionally decreased if the ego accessed SSP and had access to clean syringes (see Supplementary Information for additional details and values). Thus, each injection event with shared injection paraphernalia had a non-zero infectivity that was defined by an ego-specific transmission risk. The risk of HIV transmission was higher when a newly infected ego was in the acute phase (see Supplementary Information). Finally, initiation of ART after diagnosis decreased the base transmission probability by different rates with or without successful viral suppression. Rates of all service engagements and HIV viral suppression at baseline were based on the current uptake within the cohort (see Table S1).

### Modeled scenarios and outcomes

We assessed the relative and marginal benefits of a range of integrated harm reduction and HIV response services, increased up to the goals set by UNAID/UNODC (Figure 4). Based on the cost-of-service delivery, we assumed that services would be increased starting with the least expensive (SSP) to the most expensive (ART prescription), with MOUD and HIV testing in between. For example, a 50% goal for SSP coverage needed to be reached before a 50% goal for MOUD coverage could be reached. We considered a range of service coverage percentages in each scenario, corresponding to 50%, 75%, or 100% completion of the respective service coverage goals (Figure 4). In addition, we investigated the number of venues where these services would be deployed. We considered increased services from 1 to 12 injection venues and then 110 venues out of 110 total venues that were visited by more than 10 unique participants. Since covering 12 venues could theoretically cover >75% unique PWID in the cohort, we primarily focused on increasing venues from 1 to 12. In the model, we assumed that all individuals who frequented a venue would have an increased probability of service engagement if that venue had an expansion of that specific service; we did not incorporate correlations between engagement of one service with another.

We compared two strategies for choosing the order in which venues would be prioritized to receive all scaled-up services. For the first strategy, we ranked venues based on the venue visitor size (population-weighted approach), which was calculated as the total number of unique PWID who frequented this venue (Figure 2b). In the second strategy, we ranked venues based on geographically stratified venue population to target the most populated venue(s) that were in nonadjacent geographic clusters (spatially-stratified approach, Figure 2c). The effects of scaled-up interventions with either targeting strategy were measured as the decrease in HIV incidence in the simulated population compared to baseline, where no additional interventions were delivered. Predicted HIV incidence from our model was out of individuals who were alive in the current step of simulations. The baseline scenario assumed that service coverage stayed the same for the entire duration of the simulation at the estimated current coverage based on the proportion of visitors of a certain venue who reported access to each service. Our model was calibrated to a stable HIV prevalence and incidence within the simulated population in the baseline scenario.

## Data availability

A synthetic raw dataset and all derived datasets generated in this study[25] are available at https://doi.org/10.5281/zenodo.17971787.

## Code availability

The codes reproducing the results of this study[25] are available at https://doi.org/10.5281/zenodo.17971787.

## Author contributions

J.W., S.H.M., S.S.S., and A.W. conceived the study. A.K.S., S.M., M.S.K., G.M.L, and C.A.L. provided data. J.W. developed mathematical model and performed quantitative analysis and visualization. J.W., S.J.C, S.H.M., S.S.S., and A.W. wrote the manuscript. J.W., S.J.C., S.H.M., A.K.S., S.M., M.S.K., G.M.L, C.A.L., S.S.S., and A.W. interpreted the findings and reviewed the manuscript.

## Acknowledgements

This work was supported by National Institute on Drug Abuse grants (R01DA041736 (S.S.S.), DP1DA060602 (S.S.S.), and DP2DA056130 (S.J.C)) and National Science Foundation 2327751 (S.S.S, A.W.).

