## Supplementary Material for "Evaluating Spatially Targeted HIV Interventions and Harm Reduction Services Among People Who Inject Drugs in a High-Burden Setting"

*Model structure*

We used the exponential random graph model (ERGM) for our network simulation model structure. The probability of a tie forming between any pair of nodes was modeled with an exponential function of network covariates chosen to reflect network structure and homophily. This was informed by sociometric network data with named drug injection partnerships between people who inject drugs (PWID) within our cohort. Here we used temporal ERGM to construct a PWID network that could handle changes in population size and composition to capture the dynamic nature of the PWID injection network[1, 2].

We adapted and extended the R package EpiModel (v.2.4.0) modeling framework[1] with customizations of modules in the application programming interface (API) to build a dynamic network-based model of HIV transmission amongst a population similar to our PWID cohort in New Delhi, India. We used a temporal ERGM model structure to simulate the dynamic injection partnerships, with each node/vertex representing an individual within PWID and an edge/tie being a partnership where the two connected individuals injected drugs together in the prior 1 month. Sociometric network survey data from the “Spatial Network Study” in New Delhi, India with 2512 baseline participants (10 index participants and 2502 subsequent recruits by via a name generator network referral methodology) was used to estimate tie existence as a function of direct and derived variables from the observed injection network data. These included the total number of edges (total number of connections in the network), concurrency (proportion of nodes with more than one edge), assortative mixing (tendency for subpopulations to come into contact based on specific attributes), and edge duration (duration of partnership in months) (Table S2). Nodal covariates that would impact the mean degree included the total number of injection venues each node (individual) frequented and whether the node was living with HIV. Nodal covariates relevant to homophily in our network model included whether two nodes had the same HIV infection status and whether they frequented the same injection venue. These nodal attributes were selected for the ERGM probability function based on a previous study of the same cohort [3]. The model updated individuals within the network at each monthly time step where individuals could (1) enter the population and join the injection network, with their initial disease status reflecting the HIV prevalence of general PWID population in New Delhi, India, or (2) permanently leave the network by death (mortality rates were calculated from the confirmed and suspected death report of the cohort (Table S2)). We assumed a steady PWID population size by drawing the number of new arrivals from a Poisson distribution with the mean being the difference between the initial population size (2512) and the number of actively injecting individuals in the current time step. Network ties between existing individuals could form or dissolve in each time step. The possible formation of a tie between two nodes was based on the network terms and nodal attributes mentioned above. The dissolution of ties at each time step was modeled separately and randomly[4] by calculating the probability of dissolution of existing ties based on the average length of edge duration (Table S2).

*Population demographics and initial conditions*

We modeled a hypothetical population of ~2512 PWID with the same initial characteristics as our empirical network cohort. Each simulated individual was assigned a current age, age of first drug injection, mean number of drug injections per month, and injection venue(s) they would visit; these characteristics were directly extracted from the individual-level network data. At each time step of the simulation, each individual's number of injections in that month was determined using a Poisson distribution with lambda being their assigned injecting frequency at the start of the simulation (time zero). The initial prevalence of HIV (37%) within our simulated population at time zero was based on biospecimens data from the baseline cohort (Table S2), and we assumed that none of the infected individuals at time zero was in an acute stage. Individuals who entered the population at a later time point would also be assigned an age, monthly injection frequency, and injection venues based on distributions fitted to empirical data (Table S2).

*Injecting partnerships*

We considered sharing injection paraphernalia as the only route of HIV transmission within the simulated PWID population, since no one from our baseline cohort reported a concurrent sexual relationship with people they injected drugs with (no overlapping sexual & injection partnership). The formation of a tie between two nodes indicated a drug injection partnership, and based on our cohort data, the sum of injections done with all an ego's injection partners each month accounted for 55% (IQR = [29%, 83%]) of the total number of monthly injections reported by the ego (Table S1&S2). During injecting events with others, the risk of HIV transmission was the result of sharing injecting paraphernalia between partners. At each time point, we modeled all drug-injecting pairs originating from HIV-infected egos regardless of the alters' disease status. The distribution of these injections among an individual (ego)'s injecting partners (alters) was determined by the monthly injecting frequencies of these alters. The probability of each shared injection being assigned to an alter was proportional to the alter's injecting frequency; in other words, the more an alter injected drugs monthly, the more likely that this alter was to inject together with the ego. Between each ego-alter pair, a proportion of their injection events together had the risk of syringe sharing. Without SSP, the base probability of syringe sharing was 30% [0%, 68%] per injecting event based on survey results (Table S1, S2). Only dual partnerships were considered in the simulation, with the probabilities of each pair of injectors sharing syringes independent from each other.

*Harm reduction services*

Medication for opioid use disorder (MOUD) has been used for medically managed withdrawal from opioids[5]. We incorporated survey data reporting frequency of opiate substitution program engagement in the prior 6 months and calculated monthly probability of MOUD access for each individual (Table S1). MOUD can proportionally reduce PWID's monthly injection frequency of individuals [13]. The proportion of decrease in injection due to MOUD was calibrated to our model's baseline scenario so that the predicted HIV incidence per 100 person-years with all service coverage as status quo stays constant throughout the simulations. Individuals with access to syringe service programs (SSP) received new syringes regularly and had a proportional decrease in the likelihood of sharing syringes with others (including passing one's used syringe to others and accepting a syringe used by others) if on SSP[6]. Like MOUD, we calculated the probabilities of monthly SSP access based on the reported frequency of needle syringe exchange program engagement by our PWID cohort (Table S1). The effect of SSP was also calibrated to a constant predicted HIV incidence in the baseline scenario and was held constant. Access to one or both harm-reduction services would apply a multiplier between 0 and 1 to the number of shared injections among a drug-injecting pair, thus decreasing the probability of disease transmission if the pair was serodiscordant.

*Injection venues and venue-specific variables*

Injection venues were identified by participants from a map when asked about places where they had injected drugs in the prior 6 months. Out of 261 pre-identified areas from the survey, each participant responded 1 or 0 if they had/had not injected drugs in this area in the past 6 months; the sum of 1's a participants reported was the total number of injection venues they frequented. Visitor counts of each venue was calculated by summing all participants who reported frequenting this venue. Individual-level variables for the simulated population including 'ever been tested for HIV', 'tested for HIV in the past month', 'adherence to ART', 'prescribed ART', 'accessed SSP in the past month', 'accessed MOUD in the past month', 'proportion of injections done with others', and 'probability of syringe sharing' were venue-specific, as these values for an individual were determined by which injection venue(s) they frequented (Table S1). Venue-specific values were summarized by aggregating individual-level data of those who reported visiting a specific venue and take means. All venues with fewer than 30 visitors were summarized into ‘others’ with one set of averaged values for all the variables mentioned above. During the simulations, as we upscaled service interventions at certain injection venues, individuals who frequented these venues would have a proportionally higher probability of service access. Individuals who visited multiple venues were assigned the maximum out of the values from the venues they frequented (for example, if the venue-specific probability of monthly SSP access was 30%, 40%, and 50% for the 3 venues an individual frequented, then this individual was assigned 50% as their probability of monthly SSP access). Individuals who exclusively visited venues with fewer than 30 visitors would be assigned values from ‘others’ from Table S1.

*HIV transmission and treatment*

The initial HIV prevalence within PWID who were actively injecting was 37% based on HIV antibody titer of our baseline cohort. A newly HIV-infected individual had 1 month of acute transmission phase when the probability of infecting others was 5 times higher[7]. HIV-negative and undiagnosed HIV-positive individuals had a venue-specific monthly probability of getting tested (see Table S1 for values and the section above for description). Initiation of ART for newly diagnosed individuals was also based on a venue-specific probability and 91.5% of them reached viral suppression[8]. Once on ART, the decrease in the probability of HIV transmission was 94% for those virally suppressed and 72.8% for those who were not completely virally suppressed[9]. The probability of sustained viral suppression by adherence to ART was also venue-specific. An HIV infection event by shared injections with an HIV-infected ego was modeled by a binomial distribution; the number of trials (risky injections) and the injection probability (P(HIV)) were scaled by multiplier(s):

$$X \sim Binomial(m * (1-\mu) * (1-\sigma), \alpha * (1-\gamma) * P(HIV))$$

Where:

- m: number of injections shared with the ego

- $\mu$: proportional decrease in injection events by MOUD
- $\sigma$: proportional decrease in syringe share by SSP
- $\alpha$: fold increase in probability of HIV transmission due to acute HIV infection (5 if ego was in acute infection phase, 1 if otherwise)
- $\gamma$: decrease in probability of HIV transmission by ART treatment (0.94 if ego was on ART and virally suppressed, 0.728 if ego was on ART but not virally suppressed, 0 if ego was not on ART)

**Figure S1. Relationship between injection venue population and risky behavior/service engagement characteristics of venue visitors.**

MOUD = medication for opioid use disorder.

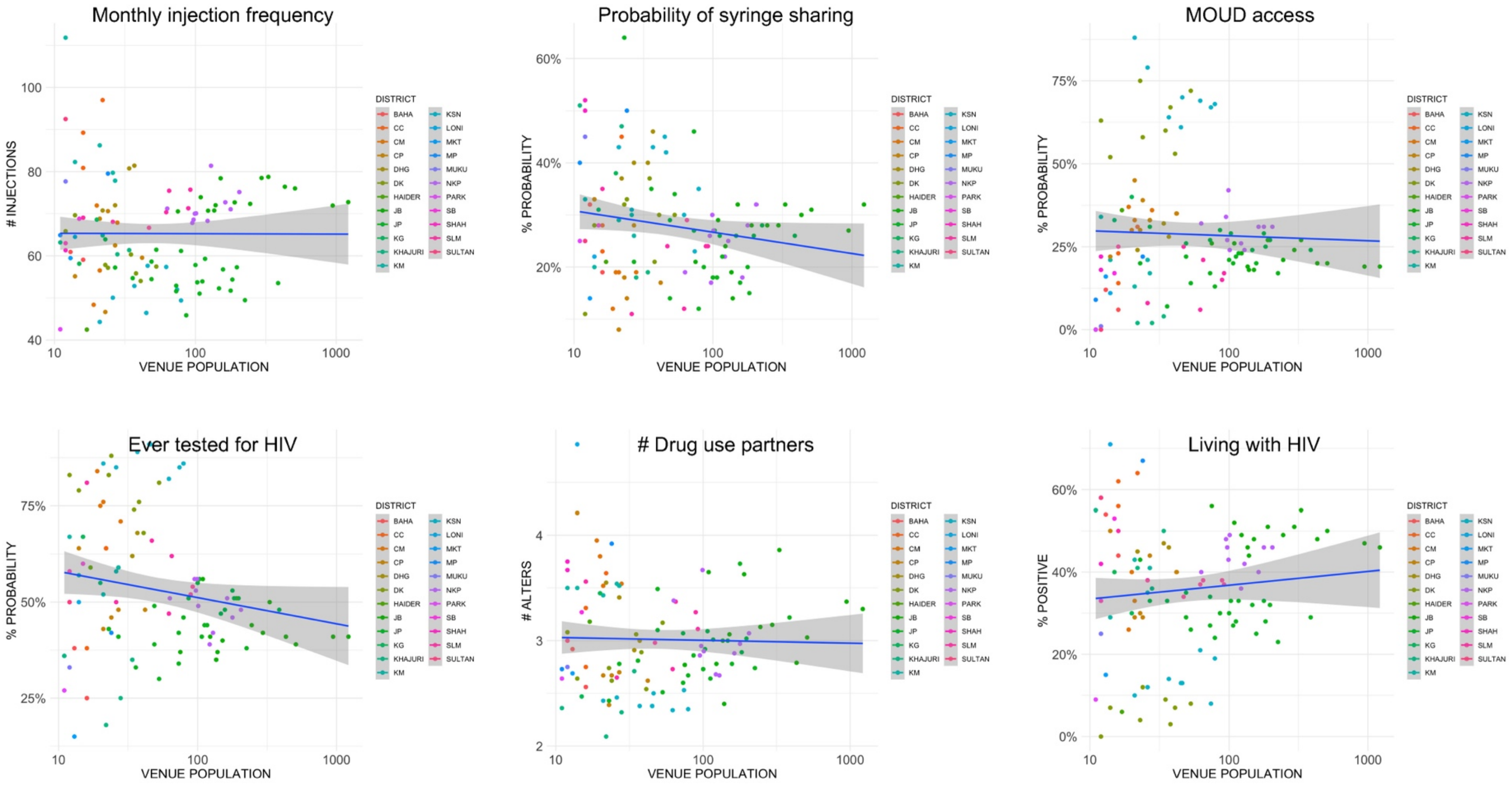

**Figure S2. Postal code map of residence of PWID surveyed within the New Delhi cohort**

Postal code regions with solid boundaries are residences of participants, and the green color scale demonstrates the number of participants residing in each postal code area. Three geographic clusters of injection venues were outlined in red.

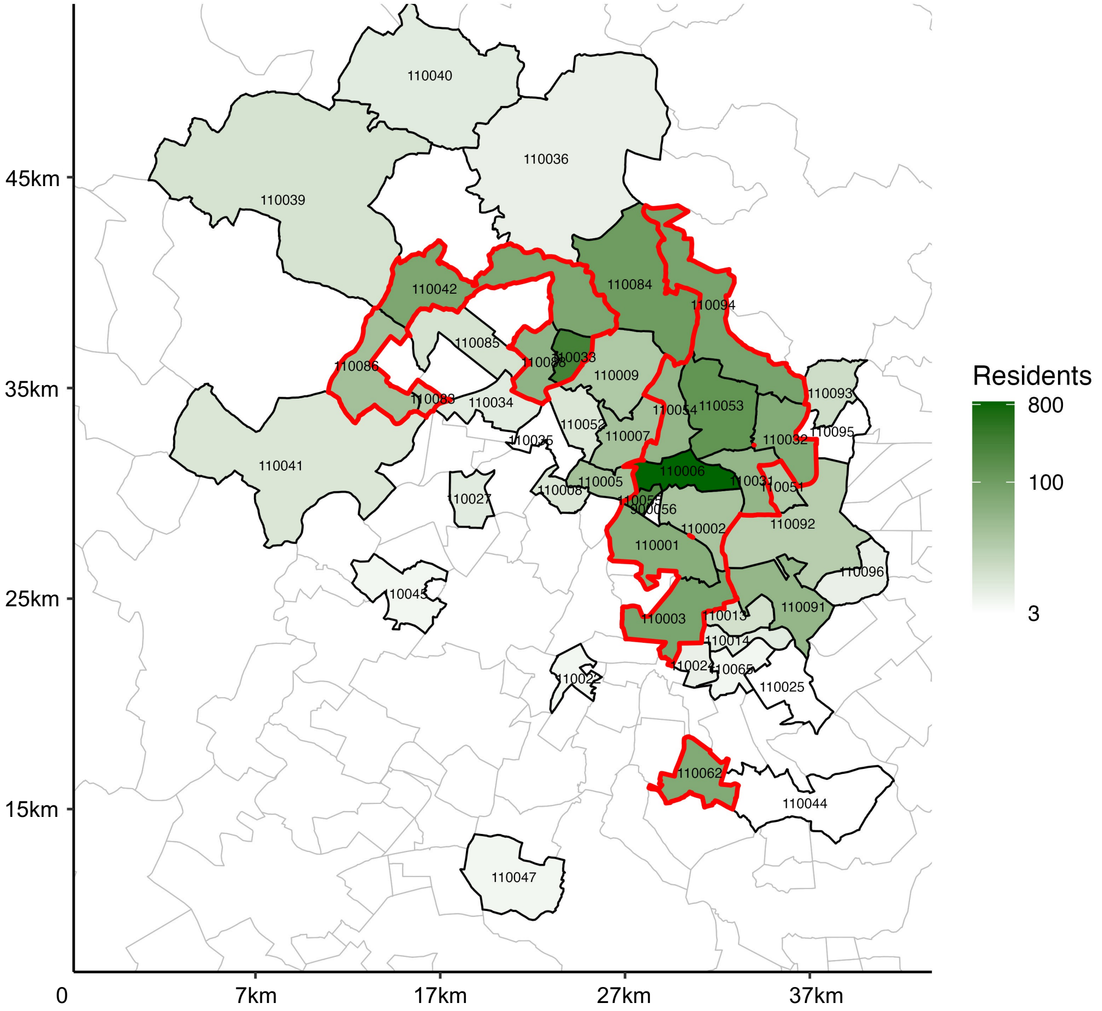

**Figure S3. Postal code areas of PWID residence covered by 3 geographic clusters of injection venues**

Map on the left is shaded based on the number of injection venues in each postal code area; 3 identified geographic clusters of injection venues are outlined in red. An arrow from each venue cluster points to a residence map of PWID surveyed within the New Delhi cohort, showing the distribution of residence areas of participants who frequented venues in this specific cluster. The color scale on the right shows percentage of residents from each postal code area frequenting this venue cluster.

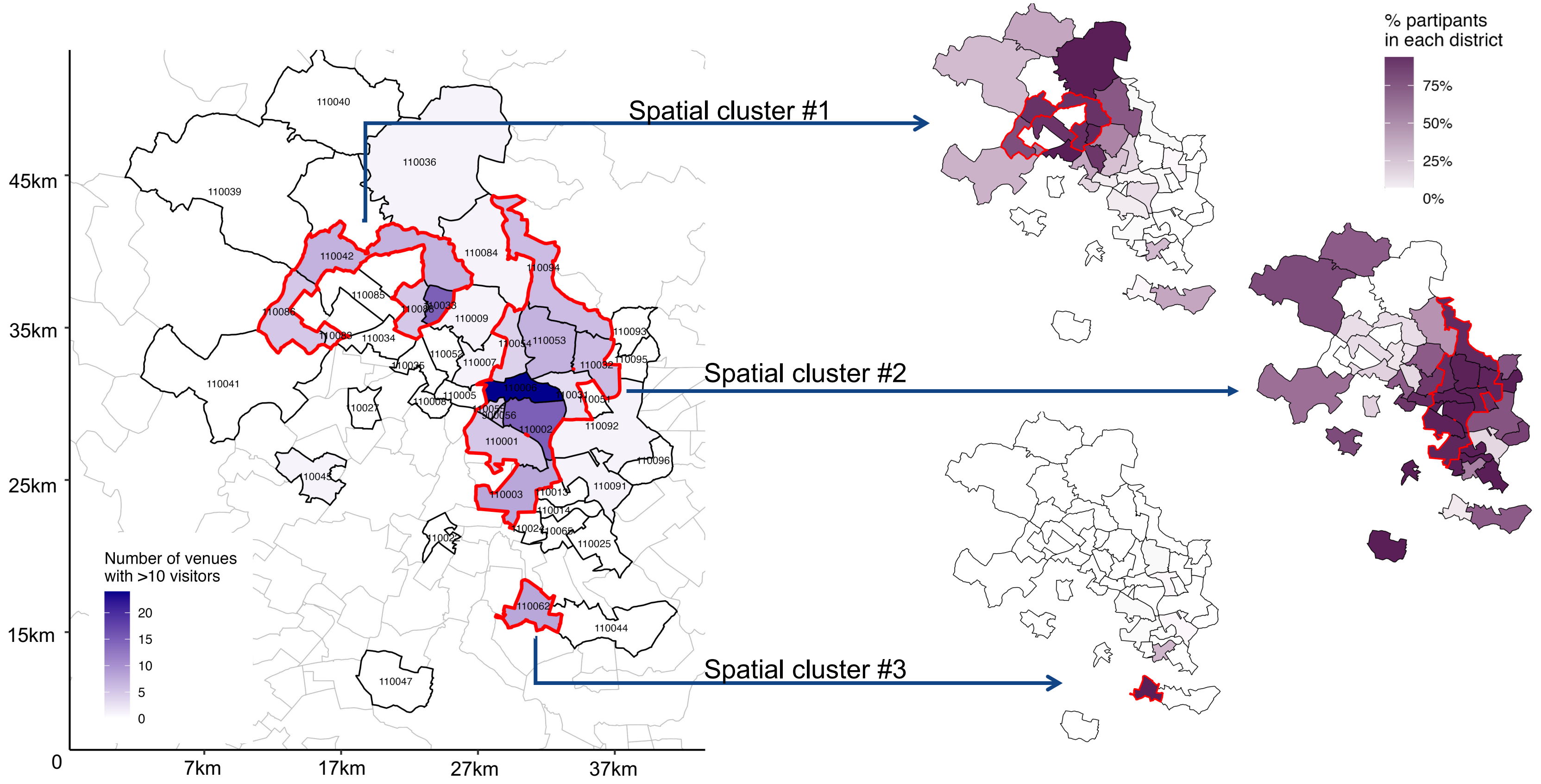

**Table S1. Venue-specific model parameters**

ART = antiretroviral therapy; MOUD = medication for opioid use disorder; SSP = syringe service program.

| Venue-level | | | Individual level | | | | | | | |
|---|---|---|---|---|---|---|---|---|---|---|
| Venue code | Visitor count | % HIV active | Proportion of injections done with others | Pr(syringe sharing) | Pr(MOUD access per month) | Pr(SSP access per month) | Pr(ART prescription) | Pr(ART adherence) | Pr(ever tested for HIV) | Pr(test for HIV per month) |
| jb1___40 | 1219 | 46% | 0.53 | 0.32 | 0.19 | 0.13 | 0.28 | 0.69 | 0.41 | 0.06 |
| jb1___39 | 945 | 47% | 0.55 | 0.27 | 0.19 | 0.12 | 0.28 | 0.61 | 0.41 | 0.05 |
| jb1___38 | 510 | 50% | 0.56 | 0.31 | 0.2 | 0.13 | 0.29 | 0.67 | 0.39 | 0.04 |
| jb1___37 | 432 | 48% | 0.55 | 0.3 | 0.2 | 0.13 | 0.29 | 0.5 | 0.41 | 0.03 |
| jp5___105 | 387 | 29% | 0.62 | 0.26 | 0.24 | 0.02 | 0.6 | 0.92 | 0.48 | 0.05 |
| jb1___41 | 330 | 55% | 0.62 | 0.32 | 0.27 | 0.21 | 0.3 | 0.73 | 0.5 | 0.05 |
| jb2___44 | 295 | 51% | 0.57 | 0.28 | 0.24 | 0.16 | 0.26 | 0.75 | 0.42 | 0.08 |
| jb2___48 | 245 | 49% | 0.62 | 0.28 | 0.21 | 0.11 | 0.33 | 0.71 | 0.44 | 0.03 |
| jp3___165 | 225 | 23% | 0.6 | 0.28 | 0.17 | 0 | 0.6 | 1 | 0.38 | 0.06 |
| nkp___52 | 205 | 46% | 0.54 | 0.32 | 0.31 | 0.08 | 0.18 | 1 | 0.48 | 0.02 |
| jp5___104 | 197 | 32% | 0.64 | 0.26 | 0.27 | 0.02 | 0.54 | 0.86 | 0.51 | 0.05 |
| jb2___47 | 190 | 51% | 0.61 | 0.28 | 0.27 | 0.17 | 0.32 | 0.5 | 0.51 | 0.02 |
| jp5___76 | 183 | 28% | 0.66 | 0.15 | 0.25 | 0.02 | 0.55 | 0.83 | 0.51 | 0.06 |
| nkp___53 | 178 | 46% | 0.55 | 0.28 | 0.31 | 0.04 | 0.38 | 0.8 | 0.46 | 0 |
| jp5___75 | 178 | 33% | 0.71 | 0.2 | 0.29 | 0.03 | 0.57 | 0.88 | 0.53 | 0.05 |
| nkp___111 | 163 | 40% | 0.54 | 0.18 | 0.31 | 0.07 | 0.4 | 0.5 | 0.51 | 0.01 |
| jp5___77 | 157 | 25% | 0.62 | 0.17 | 0.2 | 0.01 | 0.57 | 0.75 | 0.48 | 0.05 |
| jb2___45 | 151 | 48% | 0.58 | 0.32 | 0.18 | 0.12 | 0.31 | 0.5 | 0.4 | 0.1 |
| jp9___83 | 147 | 32% | 0.68 | 0.26 | 0.24 | 0.01 | 0.67 | 0.83 | 0.47 | 0.06 |
| jb1___36 | 139 | 44% | 0.58 | 0.14 | 0.18 | 0.08 | 0.33 | 0 | 0.37 | 0.06 |
| jb1___35 | 136 | 46% | 0.61 | 0.19 | 0.19 | 0.1 | 0.5 | 0.25 | 0.35 | 0.04 |
| nkp___110 | 129 | 42% | 0.49 | 0.25 | 0.24 | 0.07 | 0.43 | 1 | 0.42 | 0.02 |
| jb2___42 | 123 | 49% | 0.66 | 0.24 | 0.23 | 0.11 | 0.31 | 0.5 | 0.41 | 0.02 |
| nkp___132 | 122 | 36% | 0.48 | 0.22 | 0.26 | 0.07 | 0.25 | 0 | 0.39 | 0.02 |
| jp6___80 | 117 | 33% | 0.64 | 0.22 | 0.23 | 0.02 | 0.5 | 1 | 0.44 | 0.04 |
| jp3___136 | 112 | 28% | 0.64 | 0.26 | 0.22 | 0.01 | 0.5 | 1 | 0.44 | 0.06 |
| jb2___46 | 109 | 52% | 0.57 | 0.29 | 0.26 | 0.12 | 0.4 | 0.25 | 0.56 | 0.1 |

| | | | | | | | | | | |
|---|---|---|---|---|---|---|---|---|---|---|
| jp5___159 | 107 | 27% | 0.69 | 0.18 | 0.2 | 0 | 0.67 | 0.5 | 0.41 | 0.05 |
| jp9___82 | 103 | 33% | 0.7 | 0.27 | 0.29 | 0.02 | 0.71 | 0.8 | 0.56 | 0.05 |
| nkp___51 | 101 | 49% | 0.54 | 0.27 | 0.24 | 0.05 | 0.5 | 1 | 0.49 | 0.02 |
| jp10___85 | 100 | 30% | 0.64 | 0.18 | 0.21 | 0.01 | 0.57 | 1 | 0.55 | 0.02 |
| nkp___50 | 99 | 43% | 0.6 | 0.3 | 0.42 | 0.12 | 0.57 | 1 | 0.56 | 0 |
| nkp___134 | 97 | 46% | 0.55 | 0.17 | 0.27 | 0.08 | 0.4 | 0.5 | 0.53 | 0 |
| nkp___49 | 95 | 48% | 0.63 | 0.26 | 0.34 | 0.03 | 0.25 | 1 | 0.56 | 0 |
| slm2___33 | 92 | 37% | 0.57 | 0.24 | 0.17 | 0.08 | 0.38 | 1 | 0.54 | 0.1 |
| slm3___107 | 89 | 38% | 0.63 | 0.24 | 0.15 | 0.07 | 0.38 | 1 | 0.52 | 0.13 |
| jp5___74 | 86 | 30% | 0.71 | 0.2 | 0.3 | 0.01 | 0.8 | 0.75 | 0.51 | 0.07 |
| ksn2___62 | 79 | 19% | 0.42 | 0.35 | 0.68 | 0.06 | 0.5 | 0.33 | 0.86 | 0.09 |
| jp6___143 | 79 | 24% | 0.59 | 0.12 | 0.13 | 0 | 0 | 0.61 | 0.46 | 0.06 |
| jb2___43 | 75 | 56% | 0.64 | 0.21 | 0.26 | 0.11 | 0.2 | 0 | 0.37 | 0.04 |
| ksn2___61 | 74 | 8% | 0.44 | 0.23 | 0.67 | 0.06 | 1 | 0 | 0.85 | 0.14 |
| jp1___164 | 73 | 34% | 0.6 | 0.46 | 0.17 | 0.01 | 1 | 1 | 0.42 | 0.1 |
| jp4___166 | 73 | 27% | 0.65 | 0.27 | 0.27 | 0.02 | 0.5 | 1 | 0.34 | 0 |
| slm2___32 | 65 | 38% | 0.6 | 0.29 | 0.21 | 0.1 | 0.38 | 1 | 0.62 | 0.12 |
| nkp___109 | 63 | 40% | 0.6 | 0.19 | 0.32 | 0.07 | 0 | 0 | 0.51 | 0.03 |
| slm3___146 | 62 | 37% | 0.59 | 0.12 | 0.06 | 0.05 | 0.75 | 1 | 0.47 | 0.07 |
| ksn1___60 | 62 | 21% | 0.39 | 0.3 | 0.69 | 0.07 | 0.4 | 0.5 | 0.82 | 0.02 |
| dk2___67 | 53 | 8% | 0.47 | 0.3 | 0.72 | 0.05 | 0 | 0.5 | 0.81 | 0.07 |
| jp5___78 | 53 | 26% | 0.58 | 0.34 | 0.14 | 0 | 0.67 | 1 | 0.3 | 0.06 |
| jp6___79 | 49 | 35% | 0.65 | 0.14 | 0.22 | 0.02 | 0.75 | 1 | 0.39 | 0.05 |
| jp9___84 | 49 | 29% | 0.72 | 0.29 | 0.26 | 0 | 0 | 1 | 0.49 | 0 |
| slm2___34 | 47 | 34% | 0.68 | 0.24 | 0.25 | 0.11 | 0.75 | 1 | 0.66 | 0.03 |
| ksn1___59 | 46 | 13% | 0.4 | 0.42 | 0.7 | 0.07 | 1 | 1 | 0.91 | 0.19 |
| ksn1___58 | 45 | 13% | 0.51 | 0.45 | 0.61 | 0.05 | 0.67 | 0.5 | 0.91 | 0.17 |
| cp3___102 | 42 | 40% | 0.53 | 0.17 | 0.35 | 0.05 | 0.4 | 0 | 0.48 | 0.05 |
| dk1___65 | 41 | 7% | 0.43 | 0.3 | 0.53 | 0.01 | 0.17 | 0 | 0.68 | 0.07 |
| dk2___68 | 38 | 3% | 0.44 | 0.21 | 0.67 | 0.05 | 0.33 | 0.5 | 0.76 | 0.07 |
| dhg2___4 | 37 | 46% | 0.68 | 0.46 | 0.28 | 0.27 | 0.17 | 1 | 0.68 | 0 |
| ksn1___57 | 37 | 14% | 0.5 | 0.43 | 0.64 | 0.02 | 1 | 0 | 0.89 | 0.12 |
| jp4___160 | 36 | 33% | 0.58 | 0.35 | 0.07 | 0 | 0.67 | 1 | 0.33 | 0 |
| dk1___66 | 35 | 9% | 0.43 | 0.37 | 0.6 | 0.03 | 0.25 | 0.5 | 0.74 | 0.04 |

| | | | | | | | | | | |
|---|---|---|---|---|---|---|---|---|---|---|
| dhg1___1 | 34 | 47% | 0.62 | 0.4 | 0.32 | 0.27 | 0.17 | 1 | 0.62 | 0 |
| khajuri1___124 | 34 | 50% | 0.65 | 0.19 | 0.04 | 0.03 | 0 | 0 | 0.35 | 0 |
| others | 1013 | 28% | 0.52 | 0.32 | 0.39 | 0.05 | 0.41 | 0.6 | 0.6 | 0.1 |

**Table S2. Table of model parameters**

| Description | Mean [IQR] | Distribution | Uncertainty | Source |
|---|---|---|---|---|
| **Population** | | | | |
| Simulated population size | 2512 | | Fixed | Data |
| PWID HIV prevalence of study cohort | 37% | | Fixed | Data |
| General PWID HIV prevalence in India | 13.7% | | Fixed | [10] |
| Age (median) | 29 [22, 34] | Gamma(10.2, 0.348) | Heterogeneity | Data |
| Age of first injection (median) | 21 [18, 26] | Gamma(11.7, 0.523) | Heterogeneity | Data |
| All-cause mortality (per 1000PY) | 124.8 [82.9, 140] | Uniform(82.9, 140) | Heterogeneity | Data |
| **Network** | | | | |
| Average node degree | 2 [1, 3] | | Heterogeneity | Data |
| Concurrency | 0.711 | | Fixed | Data |
| Assortative mixing by injection venues | 0.64 [0.62, 0.66] | | Fixed | Data |
| Edge duration (month) | 18 [6, 30] | | Fixed | Data |
| **Injection behaviors** | | | | |
| Number of injections per month | 63.7 [30, 90] | Gamma(1.65, 0.026) | Heterogeneity | Data |
| Fraction of injections shared with others | 0.55 [0.29, 0.83] | | Venue-specific | Data |
| Probability of syringe sharing per injection event | 0.30 [0, 0.68] | | Venue-specific | Data |
| **HIV transmission** | | | | |
| Probability of HIV transmission per syringe sharing event | 6.3e-3 | | Fixed | [11] |

| | | | | |
|---|---|---|---|---|
| ART effect (reduction in transmission rate)<br>With viral suppression<br>Without viral suppression | <br>0.94<br>0.73 | | Fixed | [12] |
| Fold increase in infectiousness during acute infection | 5.3 | | Fixed | [7] |
| **HIV testing and treatment** | | | | |
| Probability of being tested per month | 0.065 [0.051, 0.079] | | Venue-specific | Data |
| Proportion of individuals ever tested for HIV infection | 0.48 [0.46, 0.50] | | Venue-specific | Data |
| Probability of initiating ART once diagnosed HIV+ | 0.40 [0.35, 0.44] | | Venue-specific | Data |
| Probability of ART adherence | 0.65 [0.57, 0.72] | | Venue-specific | Data |
| **Services** | | | | |
| Probability of being on MOUD per month | 0.27 [0, 0.67] | | Venue-specific | Data |
| Reduction in the probability of shared injections with MOUD | 0.36 | | Calibration | [13] |
| Probability of being on SSP per month | 0.09 [0, 0.10] | | Venue-specific | Data |
| Reduction in the probability of syringe sharing with SSP | 0.70 | | Calibration | [6] |

**Figure S4. Cumulative unique PWID covered by incrementally adding venues to target for service delivery**

Percentage of total unique individuals covered by interventions at injection venues based on three approaches: population-weighted approach and spatially-stratified approach described in the main text, and a greedy algorithm that maximize the possible unique PWID reached for each additional venue added.

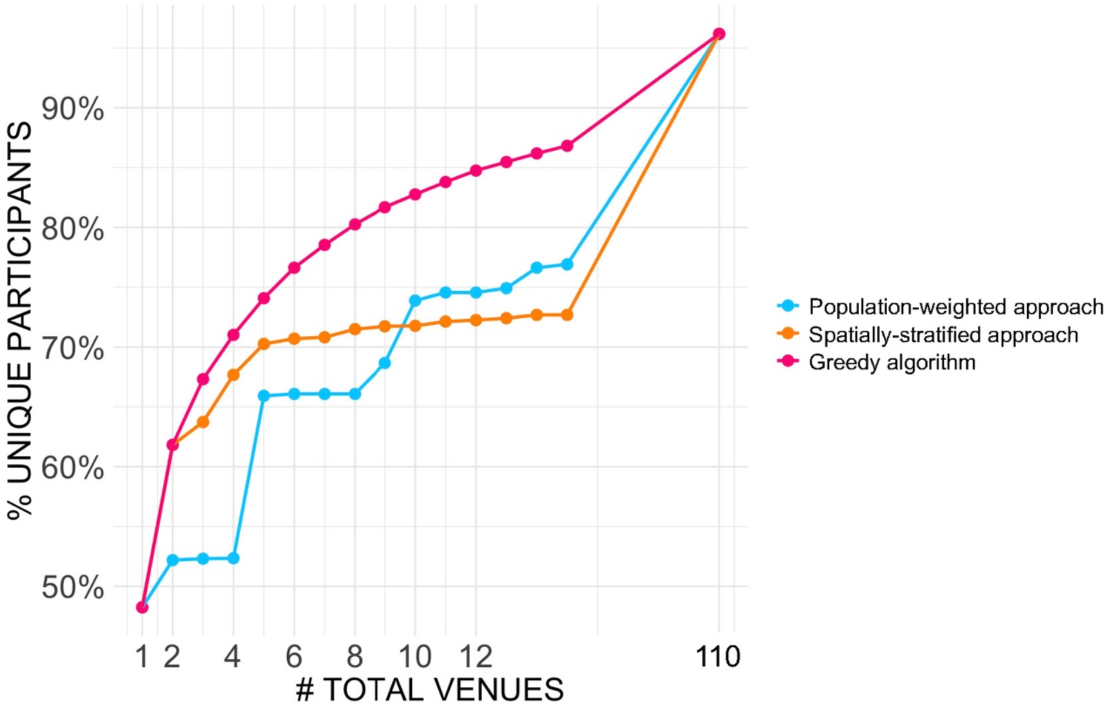

**Figure S5. Schematic description of an individual-based network model simulation of HIV transmission amongst PWID.**

MOUD = medication for opioid use disorder; SSP = syringe service program.

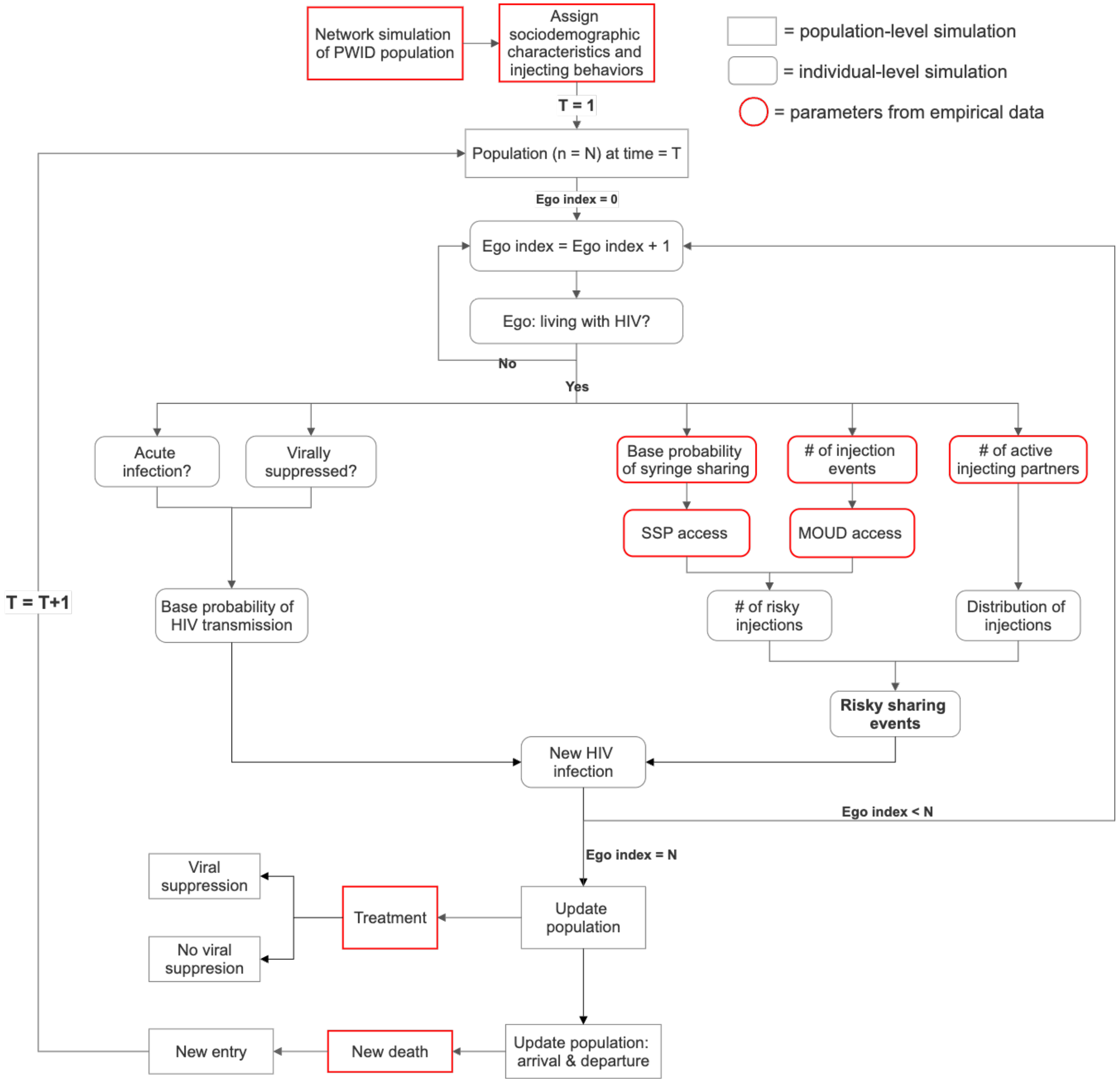

**Figure S6. MCMC diagnostics and goodness-of-fit of ERGM**

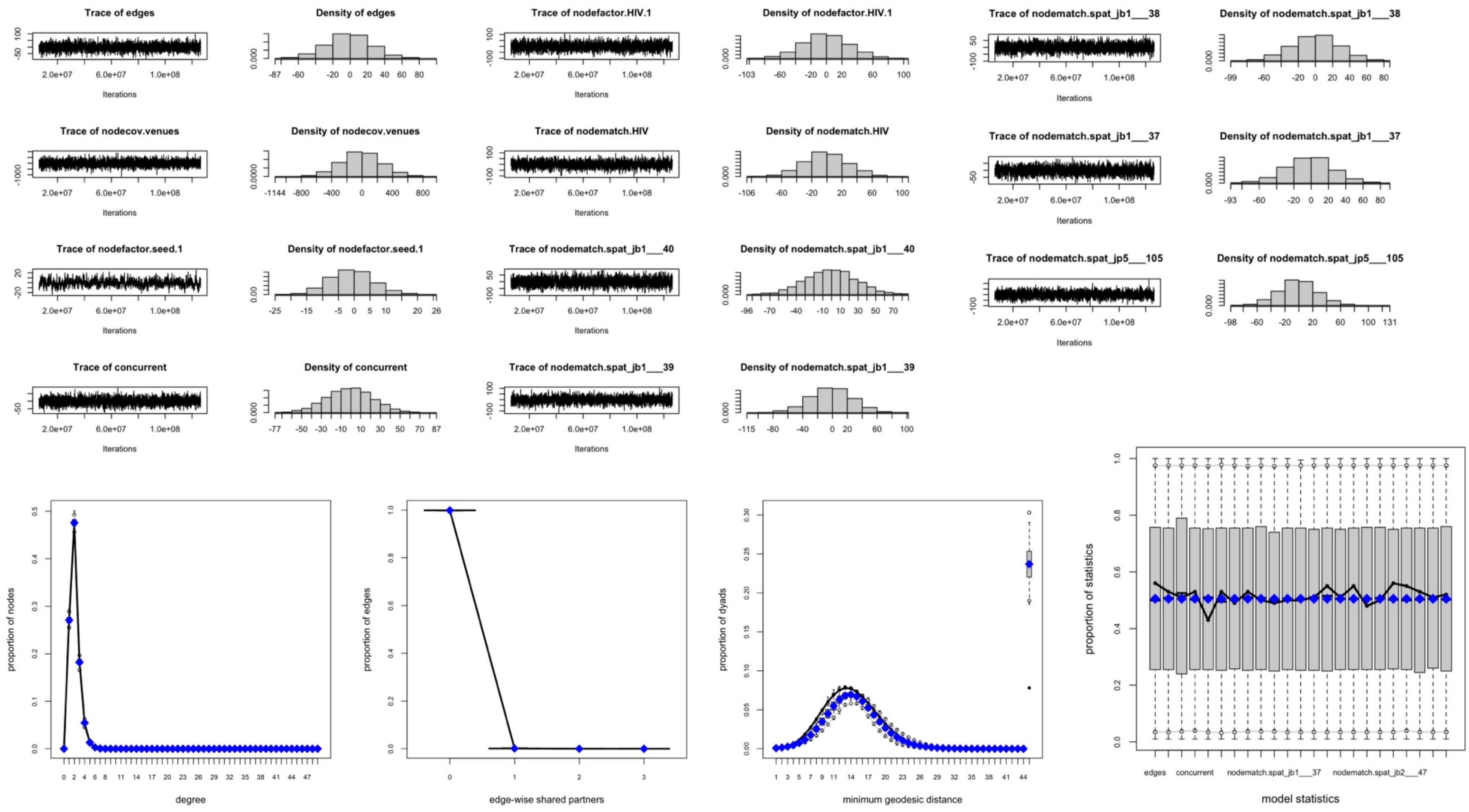

**Figure S7. Probability of SSP access two years after intervention.**

Three levels of upscaled syringe service program (SSP), medication for opioid use disorder (MOUD), HIV testing, and treatment altogether were simulated at 0 to 12 total injection venues by different venue priority ranking approaches for service deployment. Points represent mean values of 50 stochastic simulations.

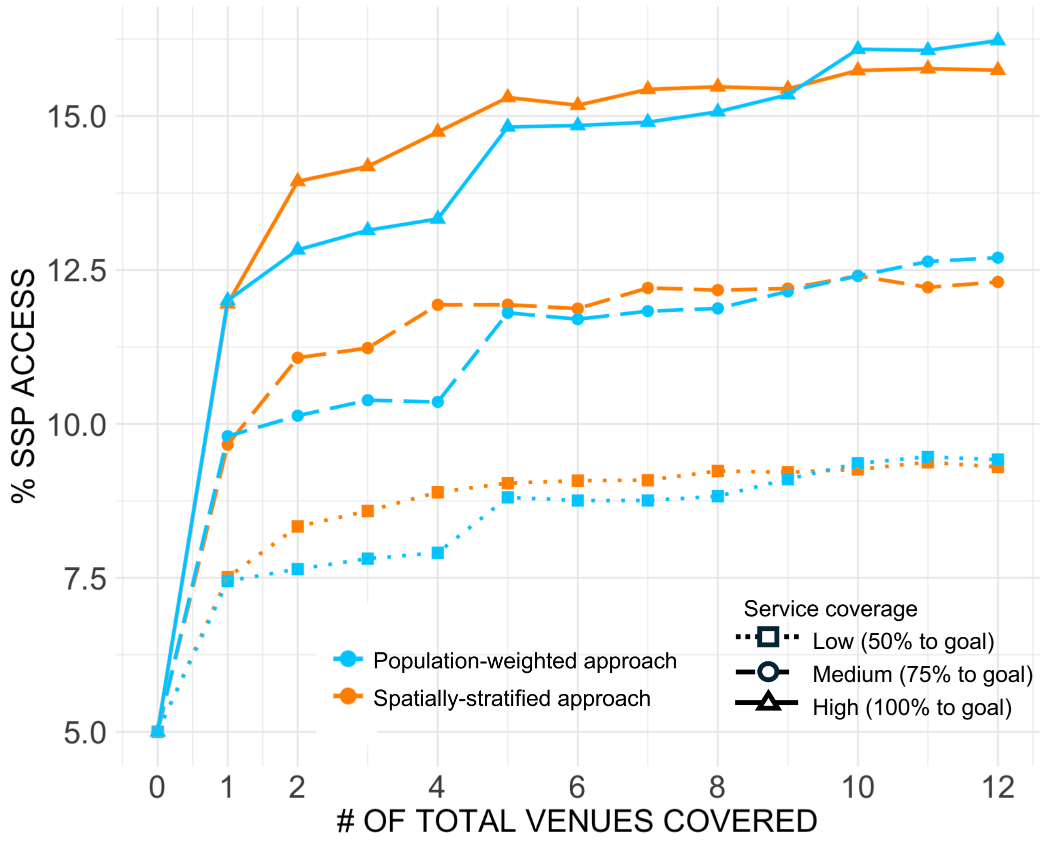

**Table S3. PWID behaviors and service access stratified by being covered by either service delivery strategy or not.**

Characteristics of a total of 2512 participants were compared by whether they reported visiting the top 12 venues prioritized by either the ‘population weighted’ or ‘spatially stratified’ service delivery strategy, and thus could be covered by the upscaled intervention simulated in the model. Two-sided Wilcoxon rank sum tests for continuous variables and two-sided Pearson’s Chi-squared tests for categorical variables were conducted. MOUD = medication for opioid use disorder; SSP = syringe service program.

| | Participants who cannot be covered by either strategy, N = 633[1] | Participants who were covered by either strategy, N = 1879[1] | p-value[2] |
|---|---|---|---|
| Age | 31 (23, 37) | 29 (22, 33) | <1e-5 |
| Age of first drug injection | 24 (18, 28) | 22 (17, 25) | <1e-5 |
| Monthly injection frequency | 54 (24, 64) | 67 (30, 90) | <1e-5 |
| Proportion of injections with other individuals | 0.51 (0.26, 0.75) | 0.56 (0.30, 0.85) | 2.7e-4 |
| Proportion of injections where syringes were shared | 0.32 (0.00, 0.80) | 0.29 (0.00, 0.65) | 0.88 |
| Ever tested for HIV | 398 (63%) | 806 (43%) | <1e-5 |
| Living with HIV | 169 (27%) | 760 (40%) | <1e-5 |
| Injection partners in the past month | 3 (1, 3) | 3 (2, 3) | 6.1e-3 |
| Number of injection venues visited | 2.6 (1.0, 3.0) | 5.0 (2.0, 7.0) | <1e-5 |
| Days accessed MOUD past 6 months | 76 (0, 180) | 39 (0, 180) | <1e-5 |
| Days accessed SSP past 6 months | 9 (0, 180) | 18 (0, 180) | 1.8e-3 |

[1] Mean (IQR); n (%)

[2] Wilcoxon rank sum test; Pearson’s Chi-squared test

**Figure S8. Cumulative unique PWID covered by incrementally adding venues to target for service delivery.**

Percentage of total unique individuals covered by interventions at injection venues based on two service expansion strategies. The x-axis showed the total number of venues covered based on either strategy, from 1 to all 110 venues with >10 individuals.

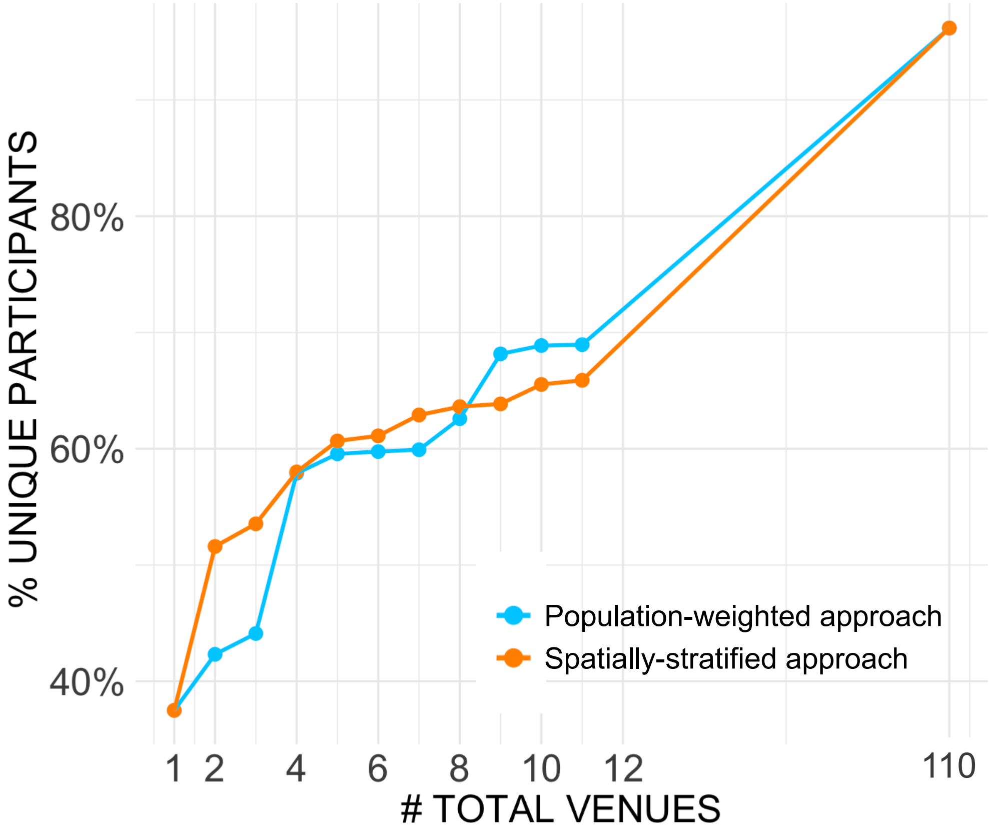

**Figure S9. HIV incidence per 100 person-years two years after intervention.**

Three levels of upscaled SSP, MOUD, HIV testing, and treatment altogether were simulated at 0 to 110 total injection venues by different venue priority ranking approaches for service deployment, dropping the most popular venue identified from our PWID cohort. Points represent mean values of 50 stochastic simulations, and error bars were upper and lower bounds of 95% confidence interval of simulated incidence values.

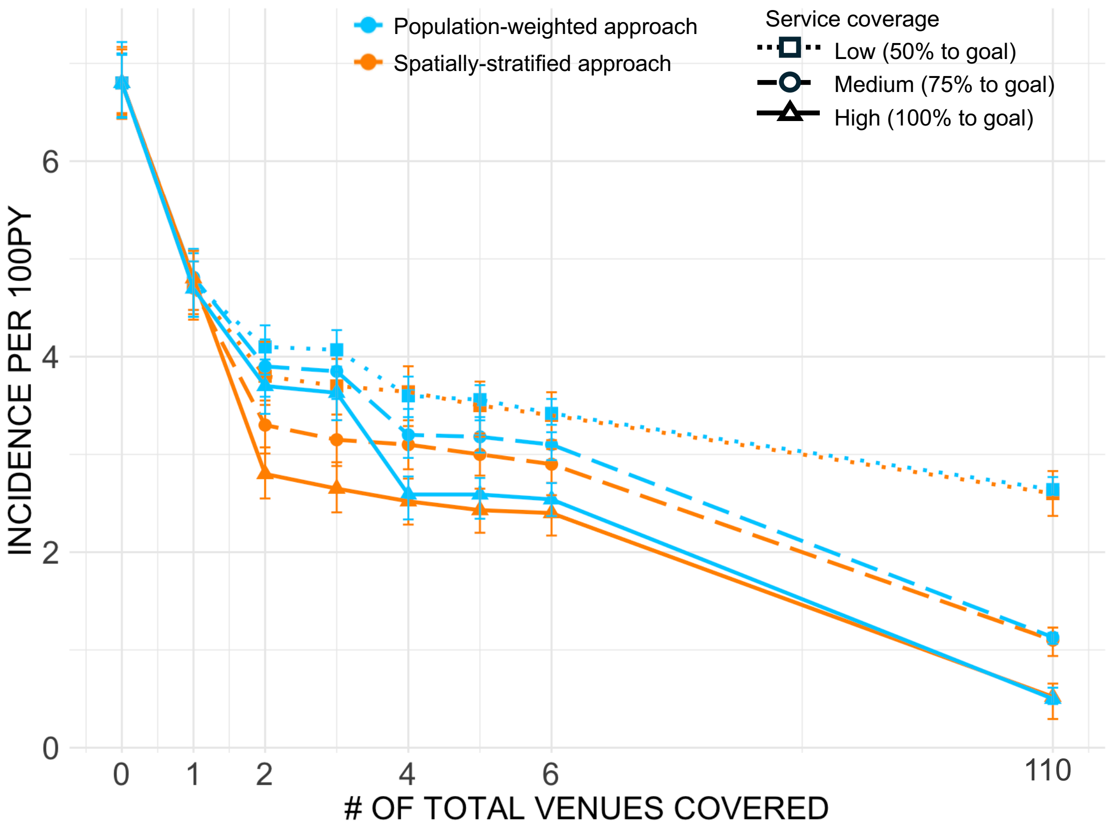
